# Developing policy to support sustainable diets in Aotearoa New Zealand with contributors

**DOI:** 10.1101/2024.10.05.24314489

**Authors:** Bruce Kidd, Hemi Enright, Christina McKerchar, Christine Cleghorn

## Abstract

Aotearoa New Zealand (Aotearoa NZ) is an example of a high-income country with high environmental impacts and health consequences associated with its food system. These impacts can be partially addressed by enabling dietary transitions to healthy and sustainable diets. The EAT-Lancet Commission proposed an internationally acceptable reference diet to promote planetary health. We aimed to work with contributors to develop policy actions to support New Zealanders to transition to healthy sustainable diets. These policies were further examined according to the World Cancer Research Fund, NOURISHING and High Level Panel of Experts (HLPE) on Food Security and Nutrition Food System frameworks. Semi-structured interviews (13) and focus groups (6) were conducted with contributors from government agencies, industry, academics, community and rural and urban tangata whenua communities. All interviews and focus groups were first transcribed using Otter.aI then reviewed by the research team. Thematic analysis was used to identify and summarise each policy. 111 policies were suggested across the NOURISHING framework policy domains: 11 (10%) in behaviour change communication; 35 (32%) in the food environment; and 65 (59%) addressing the food system. Participants spoke of behaviour change communication policies of education, awareness campaigns, and workshops such as cooking classes. Food environment policies targeted supermarkets and food retail, local food environments, and government standards and regulations including food taxes and subsidies. Policies in the food system area looked at food waste, supporting local food production and government structures and support. Contributors identified policies that advocate and support planetary health.

**Key policy insights:**

□ The majority of policies identified by contributors focused on food systems and specifically food production
□ Many contributors highlighted existing policies or initiatives already implemented locally and nationally but needed further support
□ There is limited research demonstrating the effectiveness of policies and initiatives addressing sustainable food systems in Aotearoa New Zealand
□ Funding, resources and research of existing local initiatives and policies supporting sustainable food systems in Aotearoa New Zealand and globally are urgently required

## Introduction

Food systems impose significant health and environmental impacts globally, which can be addressed by enabling dietary transitions to healthy and sustainable diets.^1–3^ Currently, food systems globally contribute to 34% of greenhouse gas (GHG) emissions from activities relating to agriculture and land use, storage, transport, packaging, processing, retail, and food consumption.^4^ Other environmental impacts include the deterioration of water resources and loss of habitat and biodiversity. In Aotearoa NZ, agriculture accounted for 50% of gross GHG emissions in 2020, due to the prominent role of the agricultural industry in the Aotearoa NZ economy.^5,6^

The EAT-Lancet Commission^2^ proposed an internationally acceptable reference diet or ‘planetary diet’ based on Rockström et al.’s^10^ planetary boundaries to promote planetary health. Six of the nine boundaries (climate change, biodiversity loss, freshwater use, interference with the global nitrogen and phosphorus cycles, and land-system change) are greatly affected by food production. The planetary diet depicts a sustainable and healthy diet as high in unprocessed plant-based foods and low in animal-based foods, consistent with the international literature.^11–13^ Much of this literature is on GHG emissions^14–17^ with less attention given to the other planetary boundaries.^18^ However, synergies between GHG emission impact of diets and impact on other planetary boundaries such as land use and water use are shown.^18,19^

The food system impacts the health of New Zealanders, with dietary risks and high body mass index accounting for the second and third highest contributors to health loss (DALYs) in 2017.^7^ Additionally, diet-related health outcomes (such as obesity) are inequitably distributed.^8^ These inequities represent a failure of policy and violate Te Tiriti o Waitangi (a treaty signed in 1840 by the Crown and tangata whenua) that includes the right to health equity.^9^ Food systems are a key area to improve planetary health through reducing emissions and the consequences of climate change on resilience and food security.

A global perspective is taken by The Commission and does not provide specific policy actions for Aotearoa NZ to support their transition to a planetary diet. Utilising a food systems approach to policy development and implementation requires strong engagement with affected contributors and communities.^20–22^ With high meat consumption and economic reliance on meat and dairy exports, Aotearoa NZ will face many challenges in achieving the necessary shifts to a planetary diet. There is a need to investigate what policies will be accepted by the Aotearoa NZ public with potential for implementation by policymakers. This includes cost-effectiveness and cultural acceptability.

Research indicates that strong engagement with contributors and policy-level support involving collaboration at different levels (such as national and local) are essential enablers for policymaking.^21–23^ This research aims to address this research gap by conducting engagement with contributors to develop policy actions specific to Aotearoa NZ. Policies were matched to the World Cancer Research Fund NOURISHING and High Level Panel of Experts (HLPE) on Food Security and Nutrition Food System frameworks to better understand their focus. We aim to describe dietary policies identified during engagement with contributors to support New Zealanders to transition towards healthy sustainable diets.

## Methods

### Positionality

The primary aim of this research was to improve health outcomes for New Zealanders with a commitment to health equity and Te Tiriti o Waitangi, and a particular focus on Māori as tangata whenua (Indigenous peoples’) of Aotearoa NZ. Bruce Kidd (he/him) and Cristina Cleghorn (she/her) are Pākehā (New Zealand European) who research food, health and sustainability and contribute to research that promotes planetary health and equity. Christina McKerchar (she/her) (Iwi affiliations are Ngāti Kahunguru, Ngāti Porou, Tūhoe) and Hemi Enright (he/him) (Iwi affiliations are Ngāpuhi, Ngāruahine, Ngāti Ruanui), are Māori health researchers in the kai (food) and food system space. We acknowledge that public health ethics of non-maleficence, social justice and an underlying philosophy of health equity guide our team.

### Engagement with contributors

Our research is part of the ‘Sustainable New Zealand Kai’ project funded by Healthier Lives He Oranga Hauora National Science Challenge (UOOX1902).^24^ Engagement protocols were finalised with the project team and advisory group. We undertook engagement among government ministries, industry representatives, academics, urban and rural tangata whenua contributors, and a community group once these protocols were finalised.

Recruitment used publicly available information to invite industry representatives (without exclusion criteria) to attend a hui (meeting) with us. In tandem with this, the authors used snowballing to recruit other contributors for interviews and focus groups. We used a semi-structured approach to explore strategies to move towards healthy, sustainable food. For organisations where only one individual participated, we conducted contributor interviews using a semi-structured interview schedule, allowing the participant to contribute beyond the stated questions. Focus groups were run by Dr Cleghorn and Dr McKerchar. 57 participants took part in one of six focus groups (with an average of seven participants in each) and 13 participants were interviewed individually.

Focus groups followed a similar semi-structured approach used for the interviews. The stated questions included asking participants how government and ministries should support New Zealanders to eat healthy and sustainable foods, what policies/interventions could help New Zealanders eat a more healthy and sustainable diet, ideas on the implementation of their policies/interventions to ensure their effectiveness, and barriers to New Zealanders consuming healthy and sustainable food.

Focus groups and interviews were recorded with contributor consent and transcribed using the software “Otter.AI”. Artificial Intelligence (AI) is increasingly used in public health research to streamline tasks and reduce human workload.^25^ AI has predominantly been developed by non-Māori without consideration of our health context in Aotearoa, Kaupapa Māori ethics or Māori Data Sovereignty, which recognises that Māori data should be subject to Māori governance.^26-28^ Otter.ai was a useful platform that allowed us to check transcripts and playback audio files while making amendments to the text. Since completing this research, the research team has reflected on the use of Otter.ai and decided to remove all audio files and transcriptions from the platform.

Notes on the specific policies were taken during interviews and cross-checked with transcriptions from recordings which were then used in thematic analysis. We used thematic analysis to identify the policies from the transcripts.^28^ The policy name was used as the code. A longer description of each policy was provided and also reviewed for key themes. When a key theme emerged across more than one policy, the policies were grouped to discuss each theme. Policies and subsequent themes were first analysed by BK and reviewed by CC and CM to confirm the final policy list and themes for the qualitative discussion.

Inclusion criteria for a policy during the extraction phase included if: the policy was from the participants’ thinking and was mentioned without a prompt from the interviewer or focus group facilitator; and/or when elaborating on a prompt from the interviewer or focus group facilitator, the participant demonstrated original thinking on the policy, e.g., providing more detail or being more specific. Policies were excluded if mentioned by a participant who then provided no further details or discussion after a prompt from the interviewer or facilitator.

Policy extraction involved noting the key information relating to the policy and further quotes from participants to provide contextual information. This included information on the policy’s scope (e.g., what population it should apply to), implementation (e.g., the resources required) or any other related information. Implementation and other related information such as barriers were discussed in more detail during subsequent interviews and focus groups with contributors.

We analysed each policy to find its matched policy domain and associated policy area (See Table 1) in the NOURISHING framework.^29^ If matched with the food system policy domain in the original NOURISHING framework (Harness food supply chain and actions across sectors to ensure coherence with health) they were subsequently matched to policy areas in the HLPE framework.^30^ Once matched, the individual policies were reviewed to identify similar policies that could be grouped to develop a final ‘unique’ list of policies. See a summary of the policy extraction process in Figure 1.

**Table 1:**
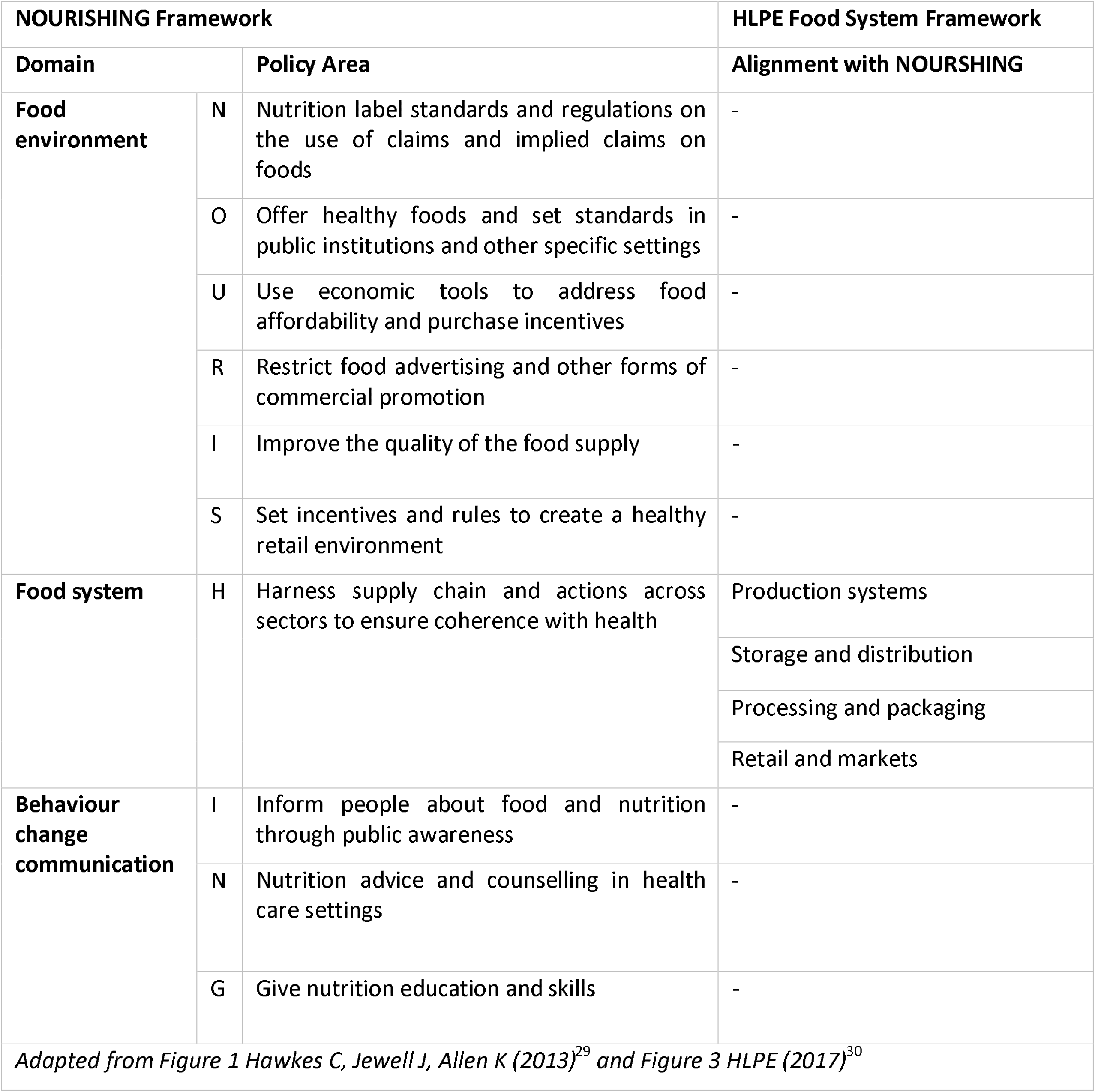
Integrating NOURISHING and HLPE Food System Frameworks.

| NOURISHING Framework |  |  | HLPE Food System Framework |
| --- | --- | --- | --- |
| Domain |  | Policy Area | Alignment with NOURSHING |
| **Food environment** | N | Nutrition label standards and regulations on the use of claims and implied claims on foods | - |
|  | O | Offer healthy foods and set standards in public institutions and other specific settings | - |
|  | U | Use economic tools to address food affordability and purchase incentives | - |
|  | R | Restrict food advertising and other forms of commercial promotion | - |
|  | I | Improve the quality of the food supply | - |
|  | S | Set incentives and rules to create a healthy retail environment | - |
| **Food system** | H | Harness supply chain and actions across sectors to ensure coherence with health | Production systems |
|  |  |  | Storage and distribution |
|  |  |  | Processing and packaging |
|  |  |  | Retail and markets |
| **Behaviour change communication** | I | Inform people about food and nutrition through public awareness | - |
|  | N | Nutrition advice and counselling in health care settings | - |
|  | G | Give nutrition education and skills | - |
| Adapted from Figure 1 Hawkes C, Jewell J, Allen K (2013)<sup>29</sup> and Figure 3 HLPE (2017)<sup>30</sup> |  |  |  |

**Figure 1:**
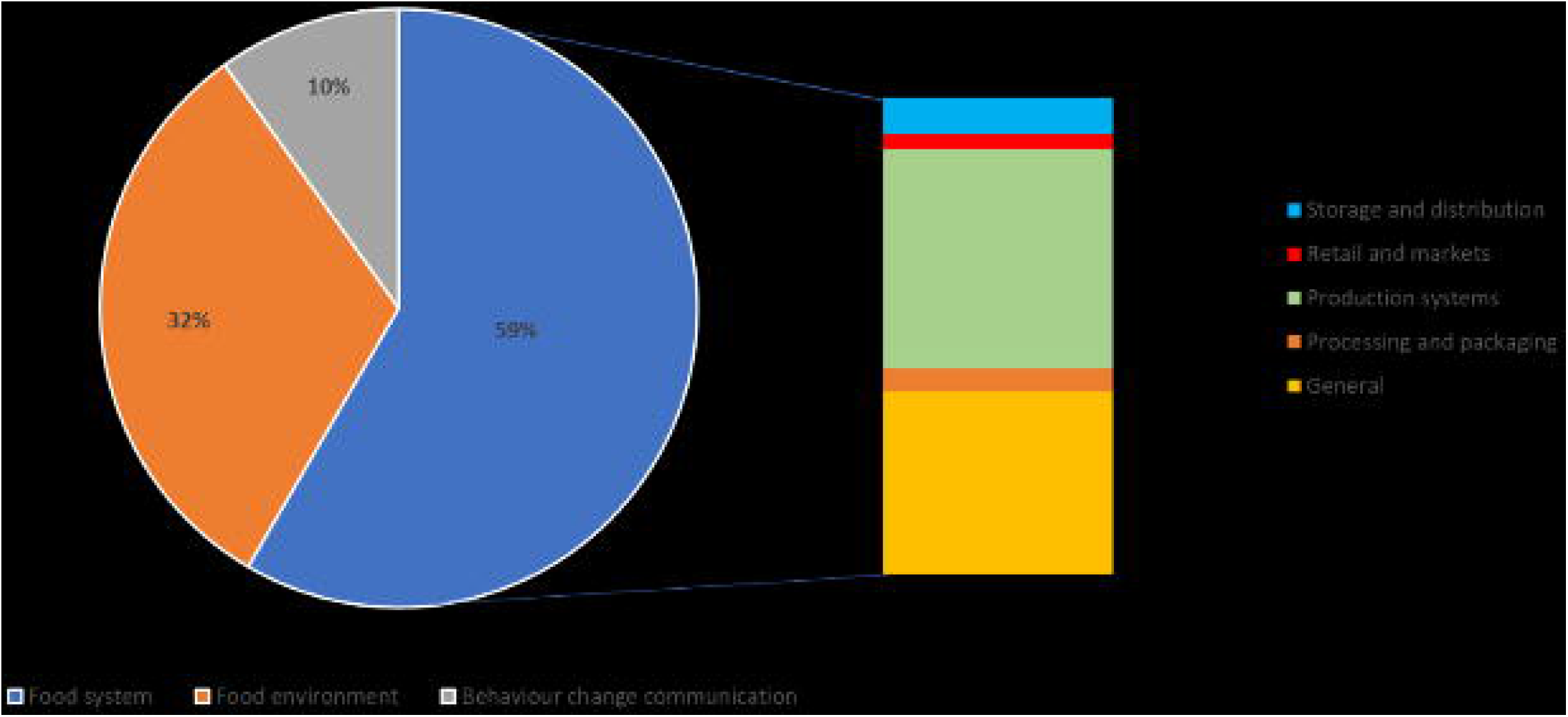
Overview of policy extraction process

The NOURISHING framework promotes healthy diets by allowing policymakers to develop suitable policy responses across the three domains. Each domain is supported by evidence demonstrating their influence on what and how populations eat.^29^ We used the NOURSHING framework as it provides a comprehensive framework detailing the different policy domains for public health nutrition.^29^ The HLPE Food System Framework supplemented NOURISHING to expand the food system policy domain, as including a strong food systems focus was important for the objectives and outcomes of this project (Table 1). The NOURSHING framework was developed by the World Cancer Research Fund (WCRF) International to provide a comprehensive policy package with key domains of action and policy areas.^29^ Microsoft Excel and Word were used to compile information on all policies and group them. When presenting quotes relating to policies’ discussed by participants, punctuation was revised and text that is not required was omitted with a ‘[…]’. A final list of policies was created using the NOURISHING and HLPE Food System Frameworks.

## Results

From the interviews and focus groups, 204 policies were identified including duplicate and similar policies. Of these 204 policies, 35 (17%) were categorised into **behaviour change communication**, 70 (34%) into **food environment**, and 99 (49%) into **food system** policy domains (**Table 5**).

Policies were then reviewed to group together these duplicates to create a list of 111 unique policies. For the complete list of unique policies and number of mentions by contributors, see ***Table 2*** in the appendix. The 111 unique policies were categorized into the following policy domains: 11 (10%) in **behaviour change communication**, 35 (32%) in the **food environment**, and 65 (59%) addressed the **food system** (Figure 2). Policies and their supporting quotes are summarised below according to the NOURISHING framework (Figure 3).

**Figure 2:**
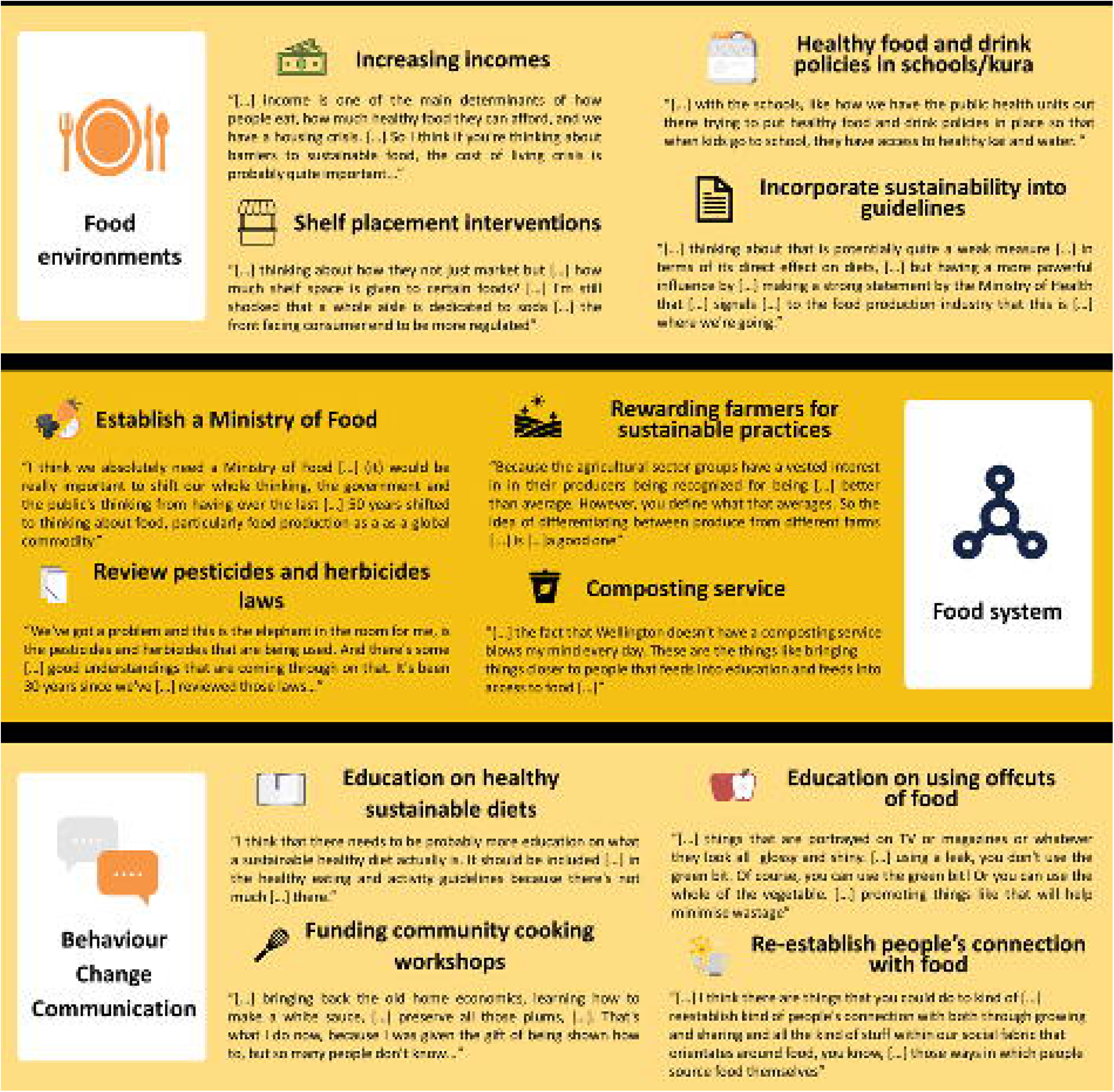
Unique policies by NOURISHING and HLPE Food System Frameworks

**Figure 3:**
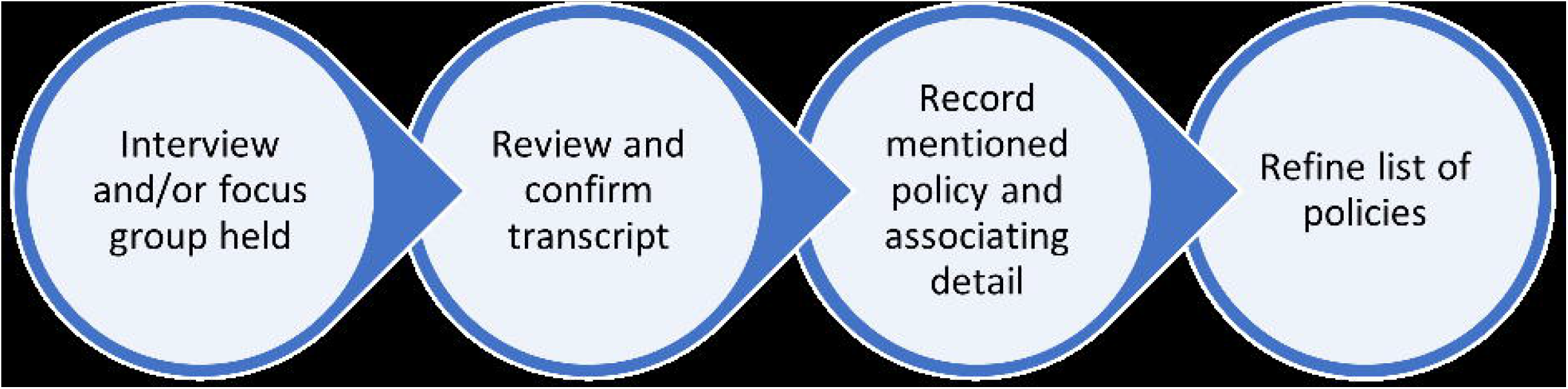
Overview of policies suggested by contributors

### Food environment

The 35 unique **food environment** policies were identified across a range of settings, including supermarkets and food retail, farming and local food environments, food industry regulations, and government standards and regulations. Policies addressing supermarkets included advertising limits on promoting unhealthy food, shelf placements interventions, promoting recipes, low-cost ‘food bags’, and local food. Contributors discussed a need to diversify products sold by supermarkets to focus on health and the environment. Spaces in supermarkets were suggested to be re-organised to provide an improved experience for families when shopping, one suggestion was supermarket playgrounds for children.

Another person, similarly identified that there was “*a missed opportunity in supermarkets. I think they can play a really critical role in influencing households and families in terms of what they purchase but how they purchase it and how they use it”*.

Other participants spoke of supermarkets’ overwhelming influence as a food environment and their dominance as a barrier to a localised, community-orientated food system. One participant said,

> *“How much profit can supermarkets be allowed to make on certain items? Thats is where I see it. Because I know how much profit they’re making. You know, like about delivering the kai to the people. It’s not about a conglomerate called Foodstuffs, making as much money as possible”*

Contributors highlighted local food environments, such as farming and gardens in different settings, including schools and local communities with healthy food and drink policies, and government standards in eating and activity guidelines. Food industry regulation was discussed, citing advertising limits, food labelling, reformulation, increasing food access, and reducing food deserts. Government standards were discussed, such as incorporating sustainability into guidelines, taxes on unhealthy foods, subsidies on healthy foods, and wealth taxes for wealth redistribution. Participants also spoke of increasing incomes through increasing wages, benefits, and introducing a living wage and universal basic income. A participant spoke of increasing wages to make healthy and sustainable food more accessible, *“[…] income is one of the main determinants of how people eat, how much healthy food they can afford, and we have a housing crisis. […] So I think if you’re thinking about barriers to sustainable food, the cost of living crisis is probably quite important*…*”*

### Food system

The 65 unique policies within the **food system** policy domain first focused on food waste including providing more compositing sites for the opportunity to compost food waste, and waste minimisation policies to remove excess materials that cannot be composted. Supporting sustainable and healthy farming practices and local food production, regulating inputs (such as fertilisers), and facilitating land use change were discussed. A participant explained the need for buy-in from farmers themselves, *“Because the agricultural sector groups have a vested interest in their producers being recognized for being […] better than average. However, you define what that average is. So the idea of differentiating between produce from different farms […] is […] a good one”*

For local food production supporting community gardens through establishing a community garden network with food hubs and growing food indoors and outdoors, enabling rural communities to barter food, access wild food sources, support local milk production and home kill, and rewarding farmers for sustainable practices were raised. The necessary infrastructure for local food production was also discussed such as implementing a nationwide permaculture policy to encourage local food production. Land use change was raised in terms of diversifying the crops grown by farmers, engaging with farmers and providing incentives.

One participant discussed shifting funds from importing and exporting food, *“you could go about it by saying […] like, we’re not going to be exporting as much and we’re not going to be able to import as much. And […] look at all these markets that are available to us […], you might have to sink some money into an indoor facility to make your citrus*”

Other system level policies raised by contributors included creating a new ministry department dedicated to food (e.g. a Ministry of Food) and reviewing legislation such as pesticides and herbicides laws. Participants spoke of the siloed and disconnected nature of how the government manages food which results in inefficiencies, *“[…] food as a concept sits over about 25 different government agencies currently and they all take a piece of the pie and wander off with it […] And I think there’s an opportunity to set up […] a government cross agency collective that is specifically about food”* This dedicated ministry could address all food-related areas, from food production to the hospitality industry. The need for a national food strategy was raised alongside key policy design principles and considerations such as food-sensitive urban design, removing industry influence, Te Tiriti o Waitangi based policy, and involving young people in decision-making.

### Behaviour change communication

Of the 11 unique policies within the **behaviour change communication** policy domain, participants predominately discussed education, public awareness campaigns and classes. Topics included food waste, education on using offcuts of food, sustainable and healthy food, gardening, cooking, land use change, farming systems, re-establishing people’s connection with food, and supporting local growers. For example, one participant explained the need for education on what a sustainable healthy diet is: *“I think that there needs to be probably more education on what a sustainable healthy diet actually is. It should be included […] in the healthy eating and activity guidelines because there’s not much […] there. It’s more about […], you need to eat this much of the stuff of food, but not really talking about where the food is coming from and taking that into consideration*.*”* There was an emphasis on linking other policies to education. Another participant discussed the links between education, access and affordability of food: *“[…] things like bringing things closer to people […] feeds into education and feeds into access to food or feed into the cost […]”*.

The importance of community engagement and a bottom-up approach to ensure buy-in of affected communities was discussed. This involved participants suggesting that the content and implementation of the policies were culturally appropriate by recognising the role of food in communities and centering their values. One participant described this as balancing cultural values with improving food waste, for example: *“And this is where we’re conflicted because we have a perception that to manaaki our whānau we need to have an abundance. [*…*]. But that needs to be balanced out…”*

## Discussion

### Main findings

Policies suggested by contributors predominately focused on the food system (especially production systems) and had a strong local and community focus, particularly on children and young people. This food system focus aligns with the large influence agriculture has on contributing to the environment compared to other sectors within Aotearoa NZ.^5,13^ Food environment policies looked at supermarkets and food retail, farming and local food environments, food industry regulations and government standards and regulations. Food system policies were directed towards food waste, local food production, supporting sustainable and healthy farming practices, facilitating land use change, and systematic policies such as within central government. The behaviour change communication policies centered education with public awareness campaigns, and classes within each policy domain.

Most policies targeting the food system appear to suggest a general agreement among our contributors on the government’s responsibility for delivering more sustainable kai in Aotearoa NZ. This aligns with previous research engaging with contributors and communities about food policy in Aotearoa NZ, for key issues of food insecurity and the right to food for Indigenous children guaranteed by the government.^31–34^ This general agreement of the states’ responsibility is particularly high among the public in Aotearoa NZ compared to other countries, with New Zealanders demonstrating the highest agreement of the government’s responsibility for food policy (70%) than any other country.^35^ The focus of food system policies and particularly production systems by the contributors in our project was similarly found in other studies involving engagement with agri-food contributors (e.g. farmers, food producers, advisors etc.) on sustainable food systems.^36,37^ Sklls and competencies considered by agri-food contributors worldwide for transitioning towards sustainable food systems included a “system perspective” which was reflected in the policies generated by our participants.^34^

The focus on production systems was reflected by a similar project in Iowa, America where sixteen organisations in the agricultural, environmental non-governmental organisations, federal and state agencies, and research organisations suggested policies for improving agro-ecosystem outcomes.^36^ Of the suggested policies, 64% were associated with land management, with examples of rewarding more sustainable farming practices and making such management practices mandatory to mitigate negative externalities.^36^ These suggested policies were similar to those raised by contributors on our project and are unsurprising given the focus on land use and dominance of food production in the Aotearoa NZ economy. Contributors in Larsen et al. (2019) identified land use planning and targeted conservation as key areas to help improve environmental outcomes with agriculture, which were similar to ideas held by our participants.^36^ Bastian and Coveney (2011) also had similar food system-level policies such as land planning policies suggested by their contributors involved in improving food security in Australia.^33^

The strong local and community focus of all interventions suggested across the three policy domains is reflected in other studies when asking contributors about their perceptions of a sustainable food system.^38^ Garcia-Gonzalez and Eakin (2019)^38^ looked at contributor perspectives in education, community building, production, distribution, policy development, waste management and processing sectors for example. Their definition of a sustainable food system included key aspects such as “local food activities”.^38^ Further, their participants discussed the role of Arizona as a large food-producing area that failed to prioritise local communities over exports. This was a similar theme among contributors in our project and their perceptions of Aotearoa NZ. As one of their participants stated, *“You can buy products at farmers markets but there needs to be policies and programs that make it possible for change to occur at a larger scale”* (p.74).^38^

Within this strong local and community focus were key policies situated in settings such as schools, community events, and local gardens, highlighting the needs of children and young people. Participants in Garcia-Gonzalez and Eakin (2019) also had this focus as “improve food access/distribution” and “edible landscape/gardening” were two top characteristics in their outline of a sustainable food system.^37^ One participant in their study explained *“Every school should have a school garden For people to be in touch with their food we need to start with the kids. I would hate to give up on adults, but there is a lost generation of people who think food needs to be fast food*.*”* (p.71).^38^

There is generally limited evidence of the policies suggested by contributors on dietary consumption and environmental outcomes such as greenhouse gas emissions.^39^ This is particularly for food system-based policies that are across sectors (agriculture, health and the environment).^39^ Further research is required to investigate these policies and resources such as funding need to be allocated to support contributors currently implementing such policies.

### Limitations

We were able to include participants and organisations from government agencies, industry, academics, community, and rural and urban tangata whenua communities. The representation of industry and urban farmers was less strong than the other contributor groups. We invited these groups to participate similarly to other contributor groups but the invitation was not accepted as commonly. We are unsure of their reasons for not participating and do not make any assumptions about why. We note this as a limitation as their representation was less considerable than the other groups. We generally had a strong response rate as 20 individuals and 7 focus groups were initially contacted to participate in the project. As with other qualitative research, it is important to acknowledge that the representativeness of the findings is limited to those who participated.

### Implications

We have engaged with contributors across various sectors to ask for their proposed solutions to help the population transition towards healthy and sustainable diets in Aotearoa NZ. To our knowledge, this is the first study to further build on the work of the EAT-Lancet Commission by generating policies that are localised to the context of Aotearoa NZ.

We found that contributors proposed a range of policies that were generally targeted at the food system level, with a particular focus on production systems. There was also a strong theme of policies proposed at the government level to assist communities with creating their own solutions.

### Future research

Research on the effectiveness of local initiatives supporting sustainable food systems is required in terms of their impacts on health outcomes (e.g. diet-related such as fruit and vegetable intake), environmental outcomes (e.g. greenhouse gas emissions and water use), and social outcomes (e.g. social cohesion). Alongside research supporting local initiatives, researchers must work with contributors and communities in developing, implementing, or enhancing new or existing policies such as community gardens and promoting plant-based food for planetary and human health.

## Conclusions

Many policies (111) were suggested by contributors on this project across community, government, industry, academia and rural and urban tangata whenua communities. Using the NOURISHING and HLPE Food System Frameworks, policies predominately focused on food systems and specifically food production. Despite limited research on their demonstrated effectiveness in the literature, this research demonstrates that there are initiatives supporting sustainable food systems that require further resources and research. Those working in sustainable food systems policy need to support contributors and communities in developing, implementing or enhancing new or existing policies to improve the health and environmental wellbeing of people in Aotearoa NZ and globally.

## Data Availability

In keeping with the consent research participants gave, the qualitative data this paper is based on is not available to share.

## Author contributions

CC and CM conceptualised the study; CC, CM, BK and HE collected and analysed the data; BK compiled the first draft of the manuscript; CC,CM, BK and HE revised the manuscript.

## Ethics

The University of Otago Ethics Committee approved this study on 13^th^ September 2021 (reference no D21/293).

## Acknowledgements

Thank you to our participants for contributing your lived experience and expertise to the project, the project advisory group, and the wider project team. This work was supported by the Healthier Lives He Oranga Hauora National Science Challenge under Grant UOOX1902.

## References

1 International Food Policy Research Institute. 2022 Global food policy report: Transforming food systems after COVID-19. Washington, DC, 2021 DOI:10.2499/9780896293991.

2 Willett W, Rockström J, Loken B, et al. Food in the Anthropocene: the EAT–Lancet Commission on healthy diets from sustainable food systems. The Lancet. 2019; 393: 447–92.

3 Food and Agriculture Organization of the United Nations, The International Fund for Agricultural Development, World Food Programme, World Health Organization. The State of Food Security and Nutrition in the World 2021. Rome: Food and Agriculture Organization of the United Nations, 2021 DOI:10.4060/cb4474en.

4 Tubiello FN, Karl K, Flammini A, et al. Pre- and post-production processes increasingly dominate greenhouse gas emissions from agri-food systems. Earth Syst Sci Data. 2022; 14: 1795–809.

5 Ministry for the Environment. New Zealand’s Greenhouse Gas Inventory 1990-2020. 2022.

6 Organisation for Economic Cooperation and Development. New Zealand. In: OECD, ed. Agricultural Policy Monitoring and Evaluation 2022: Reforming Agricultural Policies for Climate Change Mitigation. OECD, 2022: 1–652.

7 Ministry of Health. Longer, Healthier Lives: New Zealand’s Health 1990-2017. Wellington, 2020.

8 Ministry of Health. Annual Data Explorer 2021/22: New Zealand Health Survey [Data File]. 2022. https://minhealthnz.shinyapps.io/nz-health-survey-2021-22-annual-data-explorer/ (accessed Jan 6, 2023).

9 Ministry of Health. Te Tiriti o Waitangi Framework. Wellington, 2020.

10 Rockström J, Steffen W, Noone K, et al. Planetary Boundaries: Exploring the Safe Operating Space for Humanity. 2009; 14. https://about.jstor.org/terms (accessed Jan 6, 2023).

11 Springmann M, Wiebe K, Mason-D’Croz D, Sulser TB, Rayner M, Scarborough P. Health and nutritional aspects of sustainable diet strategies and their association with environmental impacts: a global modelling analysis with country-level detail. Lancet Planet Health 2018; 2: e451–61.

12 Tilman D, Clark M. Global diets link environmental sustainability and human health. Nature 2014; 515: 518–22.

13 Drew J, Cleghorn C, Macmillan A, Mizdrak A. Healthy and climate-friendly eating patterns in the New Zealand context. Environ Health Perspect 2020; 128. DOI:10.1289/EHP5996.

14 Whitmee S, Haines A, Beyrer C, et al. Safeguarding human health in the Anthropocene epoch: report of The Rockefeller Foundation–Lancet Commission on planetary health. The Lancet 2015; 386: 1973–2028.

15 Green R, Milner J, Dangour AD, et al. The potential to reduce greenhouse gas emissions in the UK through healthy and realistic dietary change. Clim Change 2015; 129: 253–65.

16 Van Dooren C, Tyszler M, Kramer GFH, Aiking H. Combining low price, low climate impact and high nutritional value in one shopping basket through diet optimization by linear programming. Sustainability 2015; 7: 12837–55.

17 Vieux F, Perignon M, Gazan R, Darmon N. Dietary changes needed to improve diet sustainability: are they similar across Europe? Eur J Clin Nutr 2018; 72: 951–60.

18 Gephart JA, Davis KF, Emery KA, Leach AM, Galloway JN, Pace ML. The environmental cost of subsistence: optimizing diets to minimize footprints. Science of the Total Environment 2016; 553: 120–7.

19 Clark MA, Springmann M, Hill J, Tilman D. Multiple health and environmental impacts of foods. Proceedings of the National Academy of Sciences 2019; 116: 23357–62.

20 Centre for Food Policy at City University of London, Results for Development. Taking a Food Systems Approach to Policymaking: A Resource for Policymakers. London, UK and Washington D.C, 2022.

21 Warren AM, Constantinides S v., Blake CE, Frongillo EA. Advancing knowledge about stakeholder engagement in multisectoral nutrition research. Glob Food Sec 2021; 29. DOI:10.1016/J.GFS.2021.100521.

22 Alliance of Bioversity, CIAT, United Nations Environment Programme, World Wildlife Fund. National and Subnational Food Systems Multi-Stakeholder Mechanisms: an assessment of experiences. 2021 https://spaces.oneplanetnetwork.org/system/files/strategy_one_planet.pdf.

23 Nguyen B, Cranney L, Bellew B, Thomas M. Implementing food environment policies at scale: What helps? what hinders? a systematic review of barriers and enablers. Int J Environ Res Public Health. 2021; 18. DOI:10.3390/ijerph181910346.

24 Healthier Lives. Sustainable New Zealand diets - Healthier Lives. University of Otago. 2022. https://healthierlives.co.nz/research/sustainable-nz-diets/ (accessed Jan 9, 2023).

25 Morley, J., Machado, C C V., Burr, C., Cowls, J., Indra, J., Taddeo, M., Floridi, L. The ethics of AI in health care: A mapping review. Social Science & Medicine. 2020; 260: 113172. DOI:10.1016/j.socscimed.2020.113172

26 Enright, J., Anderson, A., Jansen, R., Murray, J., Brewer, K., Selak, V. (2021). Iwi (tribal) data collection at a primary health care organisation in Aotearoa. JPHC. 2021; 13(1): 36–43.

27 Kukutai, T., Taylor, J. Indigenous Data Sovereignty: Toward an agenda. ANU Press. 2016; 38. http://www.jstor.org/stable/j.ctt1q1crgf.

28 Braun, V., Clarke, V. Ten fundamentals of qualitative research. In Braun & Clarke (Eds.), Successful qualitative research: a practical guide for beginners (1st Ed. pp. 19 40). SAGE Publications. 2013.

29 Hawkes C, Jewell J, Allen K. A food policy package for healthy diets and the prevention of obesity and diet-related non-communicable diseases: The NOURISHING framework. Obesity Reviews 2013; 14: 159–68.

30 High Level Panel of Experts on Food Security. Nutrition and food systems. A report by the High Level Panel of Experts on Food Security and Nutrition of the Committee on World Food Security. Rome, 2017 http://www.fao.org/cfs/cfs-hlpe.

31 Reynolds D, Mirosa M. Understandings of food insecurity in aotearoa New Zealand: Considering practitioners’ perspectives in a neoliberal context using q methodology. Sustainability (Switzerland) 2022; 14. DOI:10.3390/su14010178.

32 McKerchar C, Lacey C, Abel G, Signal L. Ensuring the right to food for indigenous children: a case study of stakeholder perspectives on policy options to ensure the rights of tamariki Māori to healthy food. Int J Equity Health 2021; 20. DOI:10.1186/s12939-021-01407-4.

33 Bastian A, Coveney J. Local evidenced-based policy options to improve food security in South Australia: The use of local knowledge in policy development. Public Health Nutr 2012; 15: 1497–502.

34 Mackay S, Gerritsen S, Sing F, Vandevijvere S, Swinburn B. Implementing healthy food environment policies in New Zealand: nine years of inaction. Health Res Policy Syst 2022; 20. DOI:10.1186/s12961-021-00809-8.

35 Pinho-Gomes AC, Booth L, Pettigrew S. Public perceptions of responsibility for recommended food policies in seven countries. Eur J Public Health 2023; 33: 299–304.

36 Larsen D, Tyndall JC, Schulte LA, Grudens-Schuck N. Exploring Stakeholder Consensus for Multiple Outcomes in Agriculture: An Iowa Case Study. Front Sustain Food Syst 2019; 3. DOI:10.3389/fsufs.2019.00110.

37 Rastorgueva N, Lindner LF, Hansen SR, Migliorini P, Knöbl CF, Flynn KM. Views of Farmers and Other Agri-Food Stakeholders on Generic Skills for Transitioning toward Sustainable Food Systems. Agronomy 2023; 13. DOI:10.3390/agronomy13020525.

38 Garcia-Gonzalez J, Eakin H. What Can Be: Stakeholder Perspectives for a Sustainable Food System. J Agric Food Syst Community Dev 2019; 1–22.

39 Reynolds AN, Mhurchu CN, Kok ZY, Cleghorn C. The neglected potential of red and processed meat replacement with alternative protein sources: Simulation modelling and systematic review. EClinicalMedicine 2023; 56. DOI:10.1016/j.eclinm.2022.101774.

