## Supplementary material for "Developing policy to support sustainable diets in Aotearoa New Zealand with contributors": Table 2; Table 3

***Table 2: List of unique policies by frameworks and number of mentions***

### Food environment

|  | **N mentions*** |
| --- | --- |
| **Policy area: Nutrition label standards and regulations on the use of claims and implied claims on foods** | |
| Eco food labelling | 2 |
| Mandating Health Star Rating system | 1 |
| Country of origin package labelling | 1 |
| Health warning labels | 1 |
| **Policy area: Offer healthy foods and set standards in public institutions and other specific settings** | |
| Garden to plate programmes in schools | 4 |
| Healthy food and drink policies in schools | 3 |
| Incorporate sustainability in MOH eating and activity guidelines | 3 |
| Access to food | 1 |
| Playgrounds for tamariki to connect with food | 1 |
| Local gardens and veggie boxes for communities | 1 |
| Reorientate shelving of products in supermarkets that compliment each other for meals | 1 |
| Mandatory sustainability guidelines for supermarkets | 1 |
| **Policy area: Use economic tools to address food affordability and purchase incentives** | |
| Food prices and taxes | 11 |
| Food taxes and subsidies | 11 |
| Increasing incomes | 5 |
| Increase incomes | 5 |
| Remove GST from food | 4 |
| Remove GST from core food | 4 |
| Increase affordability and availability of healthy and sustainable food | 2 |
| Increase affordability and availability of healthy and sustainable food | 2 |
| Wealth taxes e.g. capital gains tax | 1 |
| Low-cost food bags at supermarkets | 1 |
| Subsidise local food | 1 |
| Remove taxes on organic foods | 1 |
| Wealth taxes e.g. capital gains tax | 1 |
| Low-cost food bags at supermarkets | 1 |
| Make local food cheaper | 1 |
| Remove taxes on organic foods | 1 |
| **Policy area: Restrict food advertising and other forms of commercial promotion** | |
| Advertising limits on unhealthy food | 4 |
| **Policy area: Improve the quality of the food supply** | |
| Make sugary foods healthier | 1 |
| Diversifying food retail | 1 |
| **Policy area: Set incentives and rules to create a healthy retail environment** | |
| Government-funded school lunches | 3 |
| Address financial debt of farmers | 2 |
| Reducing food deserts | 2 |
| Food industry regulation | 2 |
| Pricing agricultural emissions | 1 |
| Encourage supermarkets to have local food | 1 |
| Shelf placement interventions for healthy and sustainable food | 1 |
| Strategies/policies for junk food and healthy food | 1 |
| Pay farmers more for organic farming | 1 |
| Healthy and sustainable 'my food bag' | 1 |
| Supermarkets promote recipes and their variations | 1 |
| Zoning limits on fast food outlets in low-income areas | 1 |

**Includes initial mention of original policy*

### Food system

|  | **N mentions*** |
| --- | --- |
| **Policy area: Harness supply chain and actions across sectors to ensure coherence with health** | |
| *Processing and packaging* | |
| Diverting food waste back into the food system | 7 |
| Composting service | 1 |
| Removing access to plastic when buying food e.g. using potato plastic instead of regular plastic | 1 |
| *Production systems* | |
| Supporting community gardens | 11 |
| Land use changes | 5 |
| Incentives for farmers to do sustainable farming practices | 2 |
| Prohibit unnecessarily destructive fishing methods | 2 |
| Regenerative agriculture policies | 2 |
| Enable rural communities to barter food | 1 |
| Freshwater policies that support Māori values | 1 |
| Phasing out synthetic nitrogen fertiliser | 1 |
| Policies to address use of chemicals, growth hormones in commerically produced animal-based products | 1 |
| Policies to support local milk production and home kill | 1 |
| Support for land owners to monitor biodiversity on their land/farms | 1 |
| Support for Māori business in youth participation and workforce | 1 |
| Support for vertical farming | 1 |
| Support to revive traditional practices such as maramataka, maara kai, mahinga kai, fishing and hunting | 1 |
| Support local growers | 1 |
| Encourage growers to diversify their crops | 1 |
| Review pesticides and herbicides laws | 1 |
| Reduce the cap on nitrogen use for farmers | 1 |
| Independent measuring systems for farmers | 1 |
| Price mechanisms for farmers to focus on quality instead of quantity | 1 |
| Community engagement with farmers | 1 |
| Promoting hedgerows on land | 1 |
| Promoting agroforestry | 1 |
| Increase access to gathered wild food sources | 1 |
| Mahinga kai | 1 |
| Nationwide permaculture policy | 1 |
| Nationwide food forests | 1 |
| Implement polyculture paddocks | 1 |
| Alternatives to imported food eg. Grow indoor citrus/tropical fruits | 1 |
| Incentivise farmers to make more sustainable choices | 1 |
| *Retail and markets* | |
| Play areas at supermarkets for kids | 1 |
| Product stewardship policies for food companies | 1 |
| *Storage and distribution* | |
| Feeding NZ first | 7 |
| Food safety reform to prevent food waste from hunted food | 2 |
| Local governments supporting community/weekend markets | 2 |
| Producer to consumer more directly | 1 |
| Shaping environments – regional council | 1 |
| *General* | |
| Food sensitive urban design | 3 |
| Establishing a Ministry of Food | 2 |
| National food strategy | 2 |
| Private investment | 2 |
| ‘Double duty’ or triple duty’ policies | 1 |
| Collect information on peoples diets | 1 |
| Current MPI priorities | 1 |
| Evidence based policy and NZ values | 1 |
| Free up funding for local communities to work on local solutions for water/pest control/ soil health etc | 1 |
| Greater engagement across sectors/ministries/ making trade offs more explicit | 1 |
| Health goals for prevention of disease | 1 |
| Health in all policies approach | 1 |
| Holistic wellbeing measure for governments | 1 |
| Improve housing affordability | 1 |
| Integrate New Zealand’s biodiversity strategy across other governmental strategies | 1 |
| Involve young people in decision making | 1 |
| Mana whenua say on local agriculture policy | 1 |
| Supporting Treasury Wellbeing Framework | 1 |
| Sustainable Food Commission | 1 |
| Te Tiriti based policy | 1 |
| True cost accounting | 1 |
| Updated national nutrition survey | 1 |
| Remove pharmaceutical lobby groups from the political sector | 1 |
| Facilitate discussions between farmers and farming organisations | 1 |
| Allowing New Zealanders to waive their rights to the health system | 1 |

**Includes initial mention of original policy*

### Behaviour change communication

|  | **N mentions*** |
| --- | --- |
| **Policy area: Inform people about food and nutrition through public awareness** | |
| Public awareness campaign about sustainable and healthy food | 7 |
| Educational support for land use changes among farmers and communities | 2 |
| Educate communities to address food waste | 2 |
| Education on gardening | 2 |
| Reestablish peoples connection with food | 1 |
| Promote natural remedies | 1 |
| **Policy area: Give nutrition education and skills** | |
| Education about sustainable and healthy food | 11 |
| Cooking classes and tailored recipes | 3 |
| Education in schools | 2 |
| Education on the impact of pharmaceuticals on farming | 1 |
| Food literacy programmes in kura, marae | 1 |

**Includes initial mention of original policy*

***Table 3: Te Reo Māori glossary***

### Te Reo Māori glossary

| **Kupu (word)** | **Definition** |
| --- | --- |
| Aotearoa | land of the long white cloud, otherwise known as New Zealand (NZ) |
| Atua | god, being of spiritual significance |
| hui | meeting |
| kai | food |
| kaupapa Māori | A collaborative approach to an issue which is uniquely Māori. |
| Kura | school |
| Mahinga kai | the practice of gathering kai, occurs within the domain of Haumia atua of uncultivated foods. |
| Mana whenua | Iwi Māori who are from this place, this area is their Tūrangawaewae (a place they stand and their ancestors stood before them and for generations to come). |
| Manaaki | to share, to give, to collaborate |
| Māori | Person who is Indigenous to Aotearoa and not Moriori |
| māra | garden |
| Māra kai | a place of storytelling, Indigenous praxis and a communal kai resource |
| Marae | the whenua that the people belong to, meet others and which their wharenui lives on. |
| Maramataka | Māori lunar calendar (Maramataka) |
| tamariki | children |
| Tangata whenua | the Indigenous people of Aotearoa (excludes Moriori) |
| Te Tiriti o Waitangi | Treaty of Waitangi written in Te Reo Māori |
| Whānau | family, collective that embraces the individual, biological and/or chosen |
